# Investigation of the Association of Acute Pancreatitis Outcomes with Social Vulnerability Indicators

**DOI:** 10.1101/2024.02.27.24302719

**Authors:** Ankit Chhoda, Anabel Liyen-Cartelle, Marco Noriega, Kelsey Anderson, Shaharyar A. Zuberi, Alana Sur, Miriam Olivares, Jill Kelly, Steven D. Freedman, Loren G. Rabinowitz, Sunil G. Sheth

**Author notes:** Corresponding author: Sunil G. Sheth, MD, AGAF, FASGE, FACG Associate Professor of Medicine, Harvard Medical School Division of Gastroenterology, Beth Israel Deaconess Medical Center 330 Brookline Ave, Rabb 423, Boston, MA 02215. Contributed equally to this manuscript (Ankit Chhoda, Anabel Liyen-Cartelle). **Author Contributorship:** AC performed the research, AC, ALC, KA, SZ, & AS collected and analyzed the data, AC & SGS designed the research study, AC & ALC wrote the paper, MO, JK, SF& LR contributed to the design of the study.

## Abstract

**BACKGROUND AND AIM:** Geospatial analyses integrate location-based sociodemographic data, offering a promising approach to investigate the impact of social determinants on acute pancreatitis (AP) outcomes. This study aimed to examine the association of social vulnerability index (SVI) and its constituent 16 attributes in 4 domains (socioeconomic status, household composition and disability, minority status and language, and housing type and transportation), with outcomes of patients with AP.

**METHODS:** This study included AP patients hospitalized between 1/1/2008 and 12/31/2018 and recorded their demographics and clinical outcomes. Physical addresses were geocoded to determine SVI, a composite variable which was ranked and divided into quartiles (I to IV: IV representing the highest vulnerability).

**RESULT:** In 824 patients [age of 53.0±10 years and 48.2% females], with 993 AP-related hospitalizations, we noted significantly higher prevalence of no/federal/state insurance ***(P<0.001*)** and racial minorities **(*P<0.001*)** in patients residing in communities with higher SVI. A non-significant trend of higher 30-day admission rate amongst patients from higher SVI regions (III/IV: 27(18.0%)/31(18.9%) *vs.* I/II: 40(13.8%)/36(16.3%); *p=*0.49) was noted. We observed a significant association of alcohol withdrawal with residence in areas with higher SVI despite adjustment for age, body mass index, and comorbidities (OR:1.62[95%CI:1.19-2.22]; *p=* 0.003). However, we observed no association of SVI with severity of AP, inpatient opioid use, length of stay, and mortality.

**CONCLUSION:** We noted significantly higher alcohol withdrawal in patients residing in areas with higher SVI ranks although AP severity and other outcomes lacked significant association with SVI. Given our findings further investigation of various social determinants of health in AP is warranted in large sized prospective studies.

## INTRODUCTION

Acute Pancreatitis (AP) is an inflammatory pancreatic disorder characterized by unpredictable outcomes and in severe cases, a substantial mortality rate of up to 20%.^1, 2^ Annually, AP accounts for a large proportion of gastrointestinal-related hospital admissions: 288,220 hospitalizations in the United States as compared to 530,855 cases of gastrointestinal bleeding. ^3^ In the United States alone, AP exerts a substantial economic impact, with annual health spending exceeding $2.5 billion and rising globally.^4^ The burden of AP is rising, as reflected by the annual percent change of 3.1% which may be attributable to improved clinical awareness of the condition, better diagnostic techniques, and exposure to attributable risk factors (e.g. alcohol use, smoking, etc.).^3,5^ Goal-directed treatment, including fluid resuscitation, is noted to be most beneficial within the first 12-24 hours of presentation.^6^

Like other health conditions such as acute coronary syndrome and reactive airway disease, overall severity and outcomes of AP may be influenced by social determinants of health (SDOH).^7, 8^ The World Health Organization defines SDOH as crucial factors that influence an individual’s well-being, as they encompass the conditions in which people are born, grow, live, work, and age.^9^ Important examples of SDOH include factors such as financial insecurity, poor access to health care, neighborhood deprivation, and ethno-racial discrimination which have previously been associated with adverse health outcomes.^10^ Many of the SDOH indicators have complex relationships that are prone to multicollinearity, making meaningful interpretation difficult. Thus, index-based approaches have emerged as proxies to assess the impact of SDOH. These approaches incorporate factors intended to capture the multidimensional nature of social determinants and their consequences for health outcomes.

Geospatial analysis has the potential to integrate location-based sociodemographic data with clinical information offering a promising approach to better understand the care of AP patients. A geospatial approach may also enable the identification of healthcare disparities, in part by making use of tools like the Social Vulnerability Index (SVI), a composite measure of vulnerability to disasters or pandemics developed by the Centers for Disease Control and Prevention (CDC).^11^ The SVI assigns each tract a raw score and a percentile rank (scored 0 to 1, with 1 representing the highest vulnerability). This multi-tiered measurement tool enables a global evaluation of a patient’s tract-level SDOH based on four thematic subdomains: (1) socioeconomic status, 2) household composition and disability, 3) minority status and language, and 4) housing type and transportation, in all comprising 16 social attributes. Each of these subdomains in turn may impact healthcare access and outcomes. The objective of this study is to assess the impact of SVI, as determined by tract-level geospatial parameters, on the health outcomes of AP patients. Through examination of various attributes determining social vulnerability, this study aims to provide insights into potential areas for future intervention and support for AP patients.

## METHODS

### Study Design and Setting

We conducted a retrospective, observational cohort study of patients diagnosed with AP and hospitalized at a tertiary care center in the Boston area between January 1, 2008, and December 31, 2018. This study was approved by the Institutional Review Board (Protocol ID: 2018P000613). This study is reported in accordance with the Strengthening the Reporting of Observational Studies in Epidemiology (STROBE) reporting guideline (**Supplement 1**). ^12^

### Study Population

Adult patients (aged 18 years or more) were identified through *International Classification of Diseases, Ninth and Tenth Revision* codes 577.1 and K85.9. ^13, 14^ Electronic health records (EHRs) were then manually reviewed to confirm the diagnosis of AP. The diagnosis of AP was based on Revised Atlanta classification (2 of the following during hospitalization: typical abdominal pain, elevation of serum lipase level three times upper limit of normal, or evidence of pancreatitis on cross-sectional imaging).^15^ Patients who were excluded were those who (1) carried a diagnosis of chronic pancreatitis or pancreaticobiliary malignancy (2) lacked physical address, thus preventing geospatial coding.

### Variable Definitions

We reviewed the EHR to collect a wide range of data, including demographic, clinical, laboratory, radiological and treatment information for patients with AP.^16, 17^ The demographic characteristics considered for each patient included age, sex, race and ethnicity, active smoking or alcohol use, etiology of AP, insurance type and address. To accurately locate each patient, we identified and geocoded their local street address. Additionally, we quantified the comorbidity burden for each participant using the Charlson Comorbidity Index (CCI), which considered pre-existing conditions such as diabetes, kidney disease, pulmonary, and cardiovascular comorbidities.^18^ The severity of presentation was assessed using the BISAP score, a scoring system that utilizes blood urea nitrogen, impaired mental status, systemic inflammatory response syndrome (SIRS), age, and the presence of a pleural effusion to predict in-hospital mortality in AP.^19, 20^ The severity of AP was based on the Revised Atlanta classification and graded as: mild AP (no local or systemic complications), moderately severe AP (transient organ failure, local complications, or exacerbation of co- morbid disease) and severe AP (characterized by persistent (>48 hours) organ failure).^15^ We also collected data on various outcomes, including the length of hospital stay for each patient, magnitude of pain upon presentation and discharge quantified by visual analog scale (VAS), opioid use quantified through morphine equivalent units (MME) required during the entire hospitalization, and the presence of local complications such as pseudocysts, peripancreatic collection, pancreatic necrosis, and splenic vein thrombosis.

Additionally, we recorded systemic complications, including renal failure, respiratory failure, bacteremia, and sepsis. We recorded the disposition of patients, including discharges to extended care facilities, the number of readmissions within 30 days, as well as mortality within one year of original hospitalization. Furthermore, we collected data on extra-pancreatic complications such as alcohol withdrawal, gastrointestinal bleeding, and delirium.

### Geocoding and Social Vulnerability Index estimation

We geocoded the residence of each patient and combined the resulting locations with the SVI metrics of the census tracts in which they are located (**Figure 1A; Supplement 2**).^11^ Developed and published by the CDC, the SVI assigns each tract a composite score which is then ranked based on percentile (ranging from 0 to 1 with 1 representing the highest vulnerability). This tool enables an evaluation of four thematic subdomains: (1) socioeconomic status, 2) household composition and disability, 3) minority status and language, and 4) housing type and transportation, in all comprising 16 social attributes (**Figure 1B).** A comparison of patient demographics and health behaviors was conducted across SVI quartiles I-IV, with quartile IV representing the highest vulnerability. Similarly, AP outcomes were also analyzed based on SVI quartiles. Maps were created using ArcGIS Pro 2.7.0 (ESRI, Red-lands, CA).

**Figure 1:**
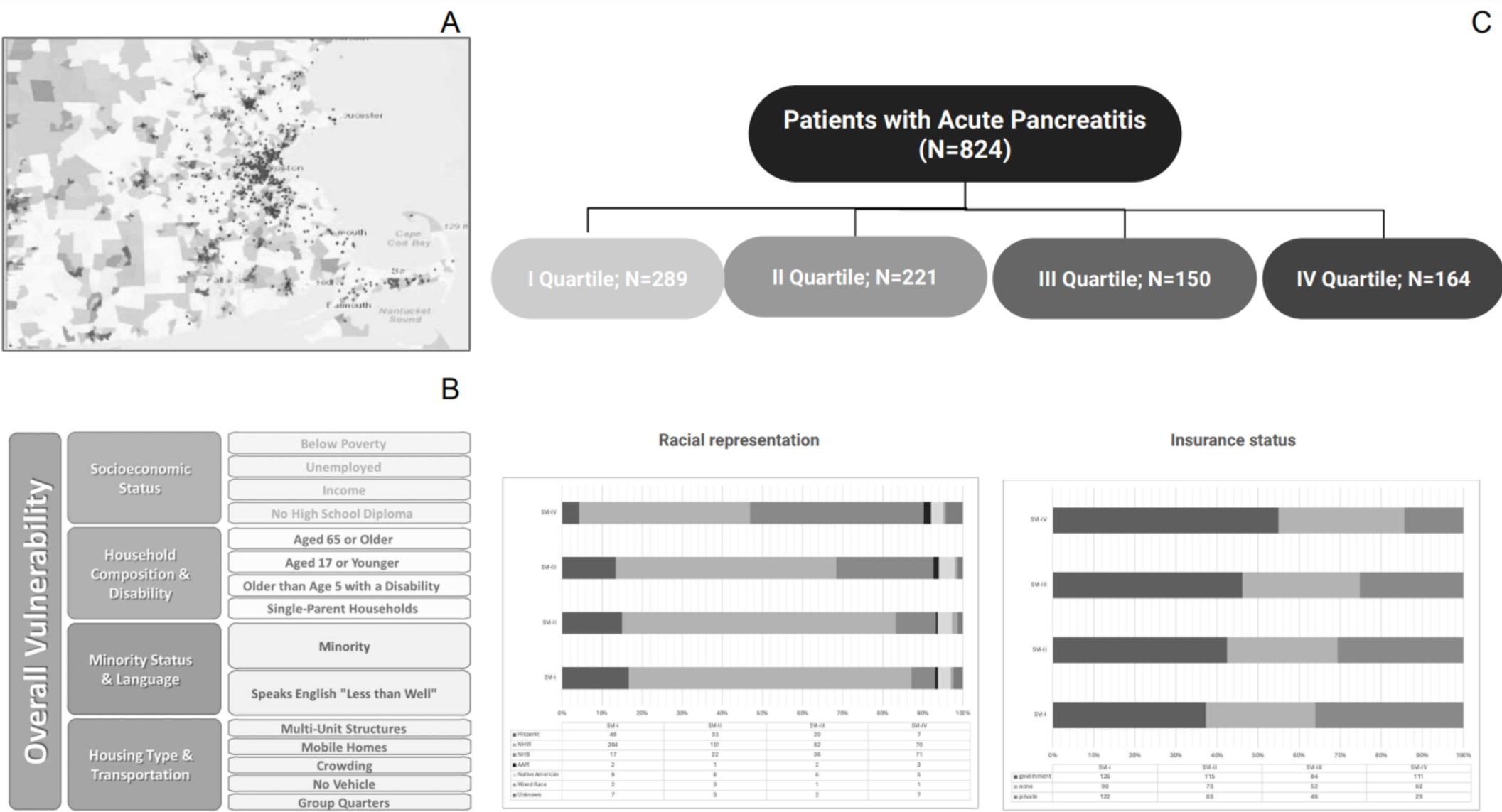
(A) Description of the patients with acute pancreatitis (AP) and areas with various social vulnerability in the greater Boston and surrounding area. (B) Description of domains and subdomains of Social Vulnerability Index (C) Description of SVI distribution and socioeconomic demographics of patients with AP.

### Statistical Analysis

Categorical data were presented as proportions, while continuous variables were reported as medians with interquartile ranges. Statistical differences in categorical variables were assessed using Chi-square tests, while continuous variables were analyzed using Kruskal-Wallis tests. AP outcome variables were the dependent variables and underwent univariate logistic regression analysis to delineate their association with SVI. Using a significance threshold of p- values < 0.1we built a multivariable logistic regression model adjusting for age, BMI, and CCI. Among variables with significant association, we analyzed the association of SVI subdomains with AP outcomes which was expressed as odds ratios (OR). The statistical analysis was carried out using STATA software (StataCorp LLC, Version 17.0, College Station, TX). A *p*-value ≤ 0.05 was considered statistically significant.

## RESULTS

The study enrolled 824 patients with AP, with a median age of 53.0(10) years and 48.18% of patients were female. There were 993 AP-related hospitalization events, for which residential locations were divided into four SVI ranking quartiles: Quartile I (n=338), Quartile II (n=271), Quartile III (n=182), and Quartile IV (n=202)

### Patient Characteristics

Baseline demographic and clinical characteristics of the study cohort and in four SVI neighborhood quartiles have been summarized in **Table 1 and** (**Figure 1C**. The study cohort exhibited ethnoracial diversity, with 146 (17.7%) non-Hispanic Black, 108(13.1%) Hispanic, and 28(3.4%) non-Hispanic Asian patients. We noted that 8 (1.0%) patients were Native Americans, 7(0.9%) were of mixed/uncategorized race, and 20(2.4%) patients lacked ethnoracial details. Most patients self-identified as non-Hispanic White (n=507; 61.7%). Neighborhoods ranked in higher SVI quartiles had a greater representation of ethnoracial minorities (*P <* 0.001), which was expected as racial and ethnic minority status is a component of the SVI. Patients residing in areas with higher SVI scores relied more on federal/state insurance or were uninsured as compared to those with private insurance (*P<*0.001; **Figure 1C**). Age and sex distribution did not differ significantly in the four residential SVI rank quartiles. Furthermore, across the SVI quartiles, there were no significant differences in self-reported healthcare behaviors, such as smoking (*p=*0.23) and alcohol use (*p=*0.23), or etiology of AP (*p=*0.20). We also noted similar comorbidity burden in the patients across all SVI quartile ranks.

**Table 1:**
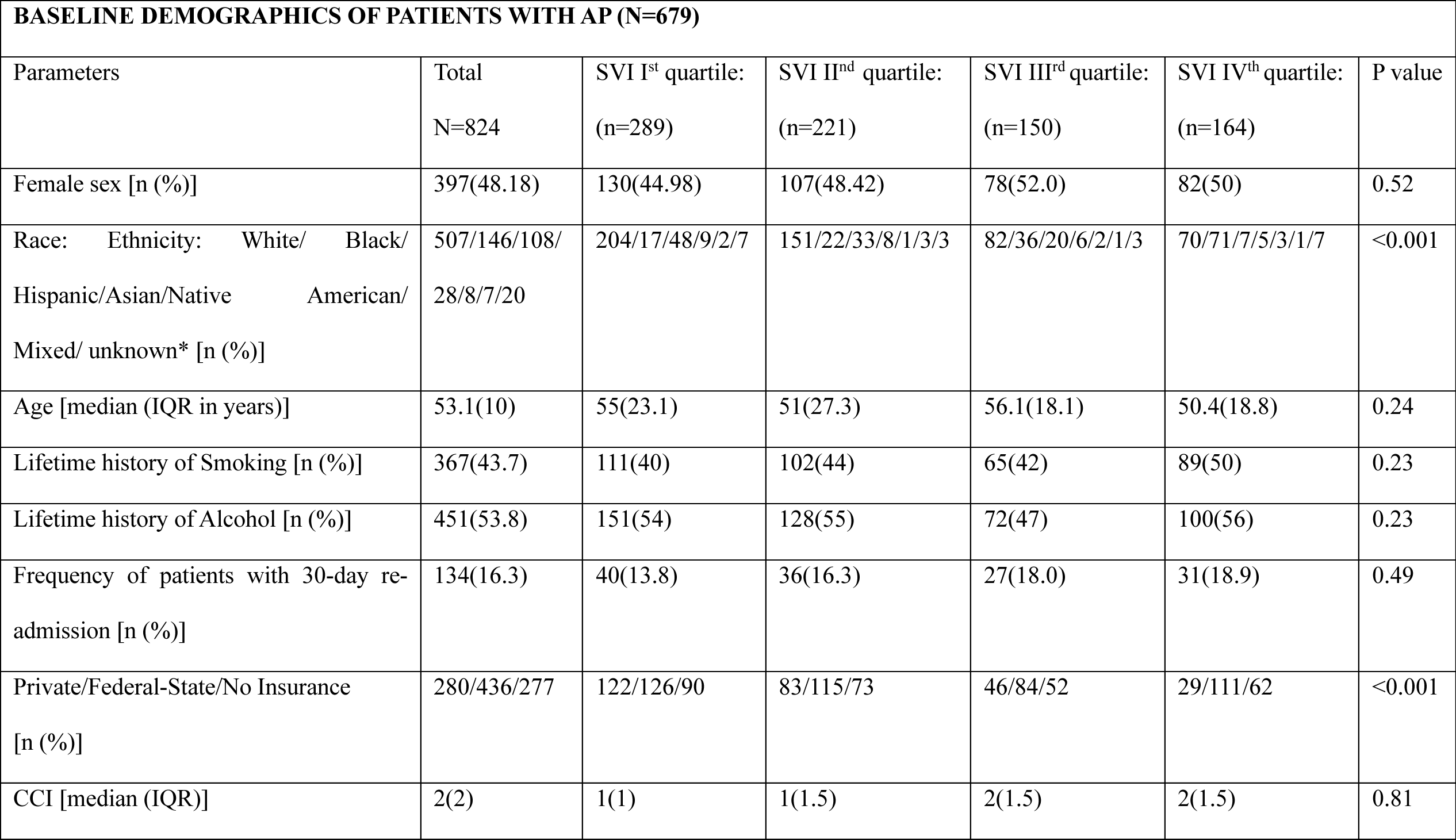
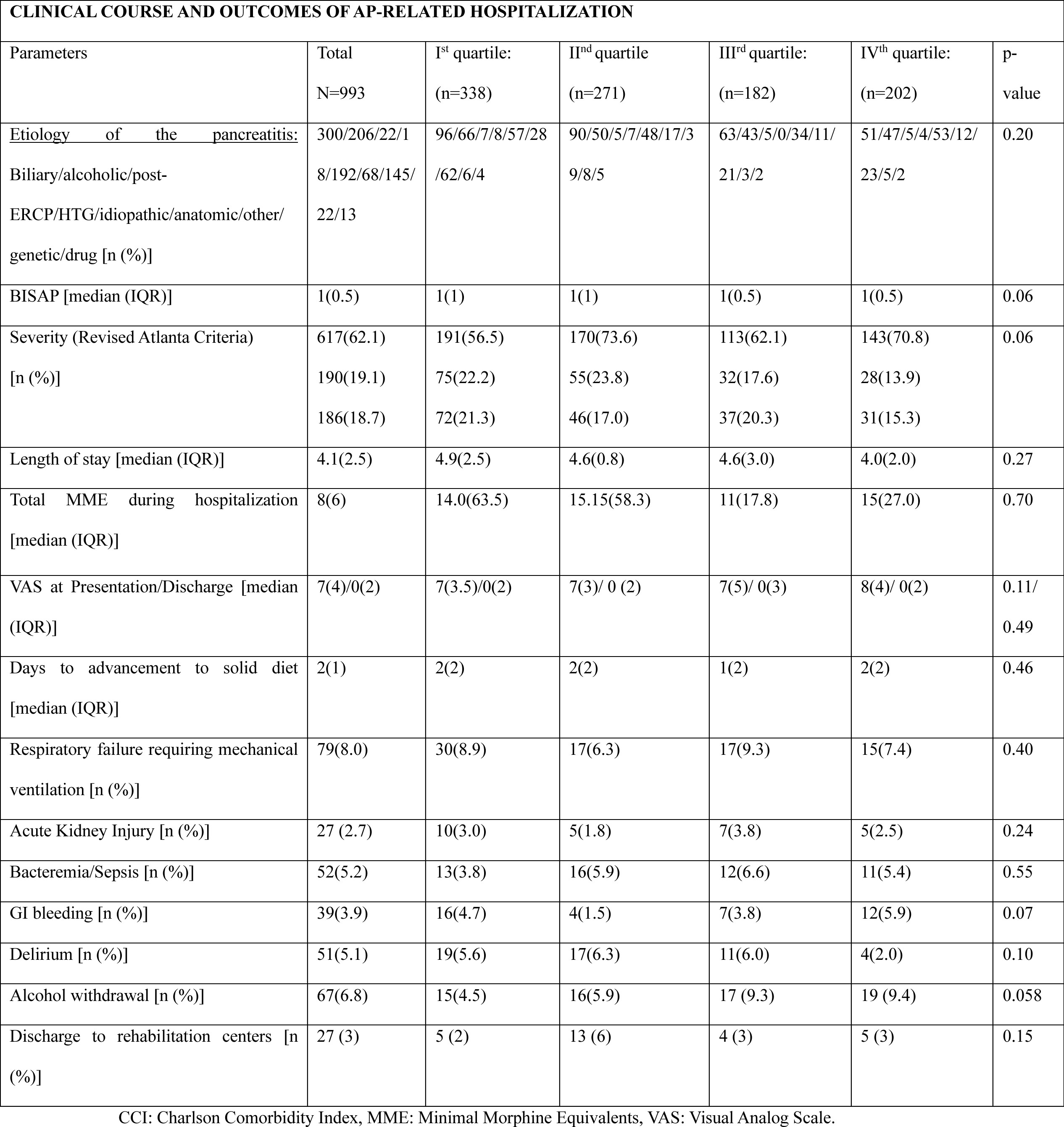
Description of baseline demographics, clinical features and outcomes in acute pancreatitis (AP)

### Clinical Course and Management

AP severity

As an indicator of the clinical severity of AP, median (interquartile range-IQR) BISAP score was 1(1),without significant difference in residential SVI quartile ranks [I:1(1); II:1(1), III:1(0.5), IV: 1(0.5); *p=*0.06]. The prevalence of moderately severe and severe AP based on revised Atlanta classification lacked significant association with SVI [I: 75(22.19%)/72(21.30%); II: 55(20.29%)/46(16.97%), III: 32(17.58%)/37(20.32%), IV: 28(13.86%)/ 31(15.34%); *p=*0.06]. The occurrence of AP-related systemic complications, including respiratory failure, acute kidney injury, and bacteremia and sepsis lacked statistical significance across the patients residing in regions with different SVI quartiles (**Table 1)**.

Management

The magnitude of pain quantified by VAS in the groups with progressive degree of social vulnerability were similar on admission [I:7(3.5), II: 7(3), III:7(5), IV:7(5); *p=* 0.11] (**Table 1)**. AP patients from all the SVI quartiles demonstrated non-significant difference in the VAS upon discharge [I:0(2), II: 0(2), III: 0(3), IV: 0(2); p=0.49]. Their opioid use quantified by MME requirements during the hospital stay lacked statistical significance [I:14.0(63.5), II:15.15(58.3.0), III:11(17.8), IV: 15(27.0); *p=*0.70] (**Table 1)**. Early enteral feeding was initiated at 2 (1) days of presentation in the overall AP population and was comparable in SVI quartiles [I:2(2), II:2(2). III:1(2), IV: 2(2); *p*=0.46]. Notably, in patients residing in higher SVI areas, the incidence of alcohol withdrawal rate was higher, and this trend approached statistical significance[I:15(4.5), II: 16(5.9), III: 17 (9.3), IV: 19 (9.4), p=0.058].

### Disposition outcome

Of the 169(17.0%) AP-related readmissions, we noted a non-significant trend of higher 30-day admission rate among patients from higher SVI regions [III/IV: 27(18.0%)/31(18.9%) *vs.* I/II: 40(13.8%)/36(16.3%); *p=*0.49] (**Table 1)**. Most patients were discharged to home, with only 27 patients requiring care at the rehabilitation center [Quartile I: 5(2%), II:13 (6%), III:4 (3%), IV:5 (3%); *p=*0.15]. One year mortality also lacked significant difference in mortality across the four residential SVI quartiles [I: 14(4.8%), II: 10(4.5%), III: 13(8.6%), IV: 14(5.4%); *p=*0.39].

### Regression Analysis

Univariate unadjusted regression analysis was performed to assess the relationship between AP outcomes and SVI quartiles, with an adjustment model made for baseline factors such as age, BMI, and comorbidities (quantified using CCI), as summarized in **Table 2**. After adjustment for age, BMI, and CCI, we noted no significant association of residence in areas with high SVI with severity of AP [0.87(95%CI: 0.75-1.02); *p*= 0.09] as well as 30-day readmission rates [1.09(95%CI: 0.90-1.33); *p*= 0.37]. The incidence of alcohol withdrawal had significant association with residence in areas with higher SVI after adjustment for age, BMI, and CCI (OR: 1.62[95%CI:1.19-2.22]; *p=*0.003; **Figure 2A).** Further analysis revealed a substantial association between alcohol withdrawal and SVI measures of socioeconomic status after adjustment for age, BMI, and CCI (OR: 7.36[95%CI:2.21-24.47]; *p=*0.001; **Figure 2B**), minority status, language (OR: 9.87[95%CI:2.55-38.30]; *p=*0.001; **Figure 2C**), housing and transportation status (6.10 [95%CI:1.44-25.79]; *p=*0.014; **Figure 2D**).

**Figure 2:**
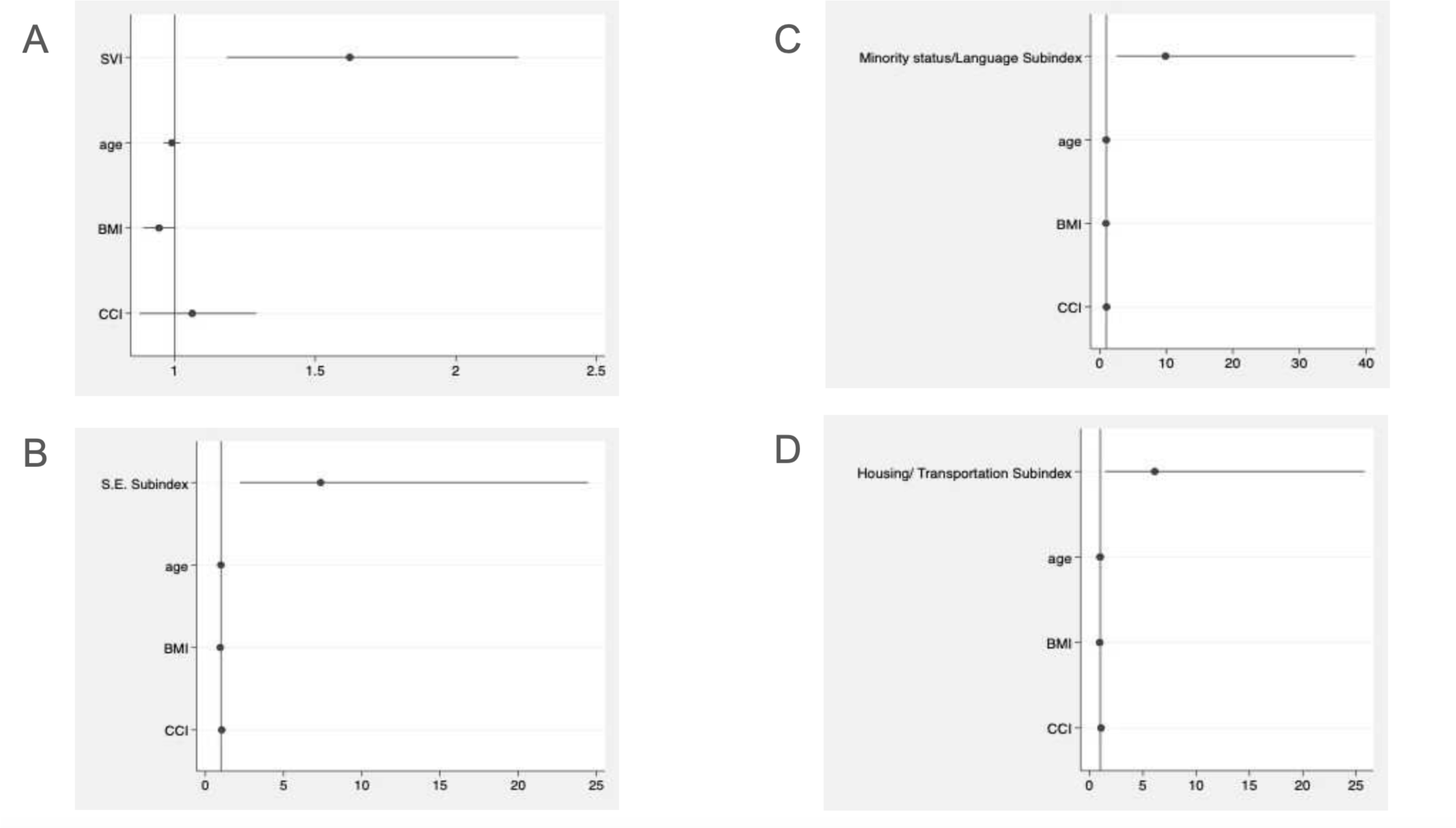
**Association of incidence of alcohol withdrawal rate with (A) SVI and (B-D) SVI subindices.**

**Table 2.** Odds ratios for the association of Social Vulnerability Index (SVI) quartiles with acute pancreatitis (AP) outcomes:

| PARAMETERS | Un-adjusted odds |  | Adjusted odds |  |
| --- | --- | --- | --- | --- |
|  | Odds Ratio | P-value | Odds Ratio | P-value |
| AP Severity (Modified Atlanta Criteria) | 0.84[95%CI: 0.76-0.95] | 0.004 | 0.87[95%CI: 0.75-1.02] | 0.09 |
| 30 days hospital readmissions | 1.13[95%CI: 0.96-1.33] | 0.13 | 1.09[95%CI: 0.90-1.33] | 0.37 |
| Mortality | 1.12[95%CI: 0.86-1.45] | 0.39 | 1.19[95%CI: 0.77-1.84] | 0.44 |
| BISAP Score | 0.83[95%CI: 0.83-0.04] | 0.001 | 0.91[95%CI: 0.79-1.04] | 0.20 |
| Length of stay | 0.94[95%CI: 0.85-1.03] | 0.85 | 0.91[95%CI: 0.81-1.02] | 0.12 |
| Alcohol withdrawal | 1.33[95%CI:1.07-1.65] | 0.01 | 1.62[95%CI:1.19-2.22] | 0.003 |
| Association of AP Outcomes with Thematic Subindices |  |  |  |  |
| Alcohol withdrawal |  |  |  |  |
| Socioeconomic status subindex | 3.14[95%CI:1.35-7.25] | 0.007 | 7.36[95%CI:2.21-24.47] | 0.001 |
| Household and disability subindex | 1.24[95%CI:0.56-2.76] | 0.60 | 1.92[95%CI:0.62-5.91] | 0.26 |
| Minority status and language subindex | 3.48[95%CI:1.41-8.59] | 0.007 | 9.87[95%CI:2.55-38.30] | 0.001 |
| Housing and transportation subindex | 3.14[95%CI:1.22-8.13] | 0.018 | 6.10 [95%CI:1.44-25.79] | 0.014 |

## DISCUSSION

The morbidity of patients with AP, its complications, and overall mortality emphasize the need for conducting comprehensive SDOH-based geospatial investigation.^1, 2^ In this study, we employ a novel approach of geocoding and stratifying patients based on the SVI scores and ranked quartiles of their residential locations, a validated SDOH grading tool, to assess unmeasured factors which may have an impact on patients hospitalized with AP. ^11^ Across SVI quartiles, we evaluated geospatial disparities in clinical outcomes including disease severity, management, disposition, and hospital readmissions.

In this retrospective cohort study, we noted that severity of clinical presentation lacked significant association with residence in communities with social vulnerability, especially after adjusting for confounders (*p=*0.09). We observed no significant difference in the local and systemic complications (e.g., respiratory failure, acute kidney injury, and bacteremia) mortality, length of hospital stays, and rehabilitative requirements across SVI quartiles, but a trend towards increased 30-day readmissions in the higher SVI quartiles. However, we observed higher odds of alcohol withdrawal among patients with residing in communities with higher social vulnerability. A significant association of alcohol withdrawal was noted with subdomains of SVI i.e. socioeconomic status, underrepresented minority status, language, housing and transportation status. This finding is consistent with prior reporting of higher community prevalence of alcohol withdrawal syndrome in single male patients in the lowest socioeconomic strata.^21,22^

Additionally, the subjective nature of assessing alcohol withdrawal (Clinical Institute Withdrawal Assessment protocol) can impede timely identification of this entity among patients with language barriers, further amplifying this problem.^23, 24^ The impact of the physical environment and sociodemographic data based on the address of the patients with AP has previously been investigated in national electronic healthcare record linkage cohort studies. In the UK, the Social Deprivation Index, a multimodal marker involving seven demographic factors (poverty, education, single-parent households, rented housing units, overcrowding, transport, unemployment), has been studied when considering AP outcomes.^25^ SVI, by comparison, may enable better capture of heterogeneity in health care coverage and geospatial/physical infrastructure systems which may impact AP outcomes, as it includes additional parameters.

Our study is strengthened by a unique design for determining clinical outcomes of AP patients and well-annotated clinical data. We relied on a manual review of individual charts, thus having high certainty of AP diagnosis. We also geocoded the exact locations of each patient, which would not be possible using publicly available databases due to privacy issues. Our conservative analysis investigated each trend with regression models which were adjusted for demographics to reduce confounding. Aggregate geospatial socioeconomic variables have been compared with individual-level data, and were confirmed to have internal validity.^26^ This was demonstrated through higher minority representation and percentage of uninsured/reliant on federal insurance in the patients with higher social vulnerability. This was done since geographical aggregation of AP patients may predispose such studies to an ecological bias.^27,26^ Although we used insurance status, absence of individual socioeconomic data such as annual income constrained our ability to account for this ecological fallacy.

The study is limited by its retrospective design and reliance on participants’ physical address, which fails to capture change in residence or migration. Geocoding residential address also limits assessment of homeless population which may be more at risk of adverse health outcomes. ^28, 29^ The study also lacks details on alcohol drinking behavior and nutritional assessment of our patients, thus limiting the inference of its impact on the trend seen for alcohol withdrawal. SVI though beneficial is not perfect. It is summation of various social attributes akin to principal component analysis and may have some critical shortcomings regarding theoretical and internal consistency.^30^

Also, it was not unexpected that in our study AP patients residing in more socially vulnerable communities did not have worse outcomes during their hospitalization, as the care provided to all is completely based on evidence-based guidelines and decision tools, as previously reported.^31^ However, as noted, alcohol withdrawal rates were higher in these patients, as this is dependent on drinking patterns outside the control of medical care givers. Moreover, being aware of patients being discharged to areas with higher social vulnerability may be particularly important for their recovery process. While we recommend careful consideration of the factors elevating community SVI scores in optimizing discharge planning and follow-up strategies, we caution against oversimplified applications of SVI in risk reduction efforts or healthcare policy aimed at addressing social determinants of health.^30^

In conclusion, this study highlights the importance of geospatial analyses in investigating the impact of social determinants on AP outcomes. SVI can be used to identify sociocultural, economic, and institutional processes that shape socioeconomic differentials in the management of AP. Communities with higher social vulnerability, as indicated by their placement in higher SVI quartiles, exhibited elevated rates of alcohol withdrawal. The integration of geospatial considerations into future AP research can contribute to a more holistic approach to the disease, enable investigations into SDOH that may influence AP outcomes, and provide opportunities for more tailored public health interventions.

## Data Availability

All data produced in the present study are available upon reasonable request to the authors

---

**Supplement 1:**
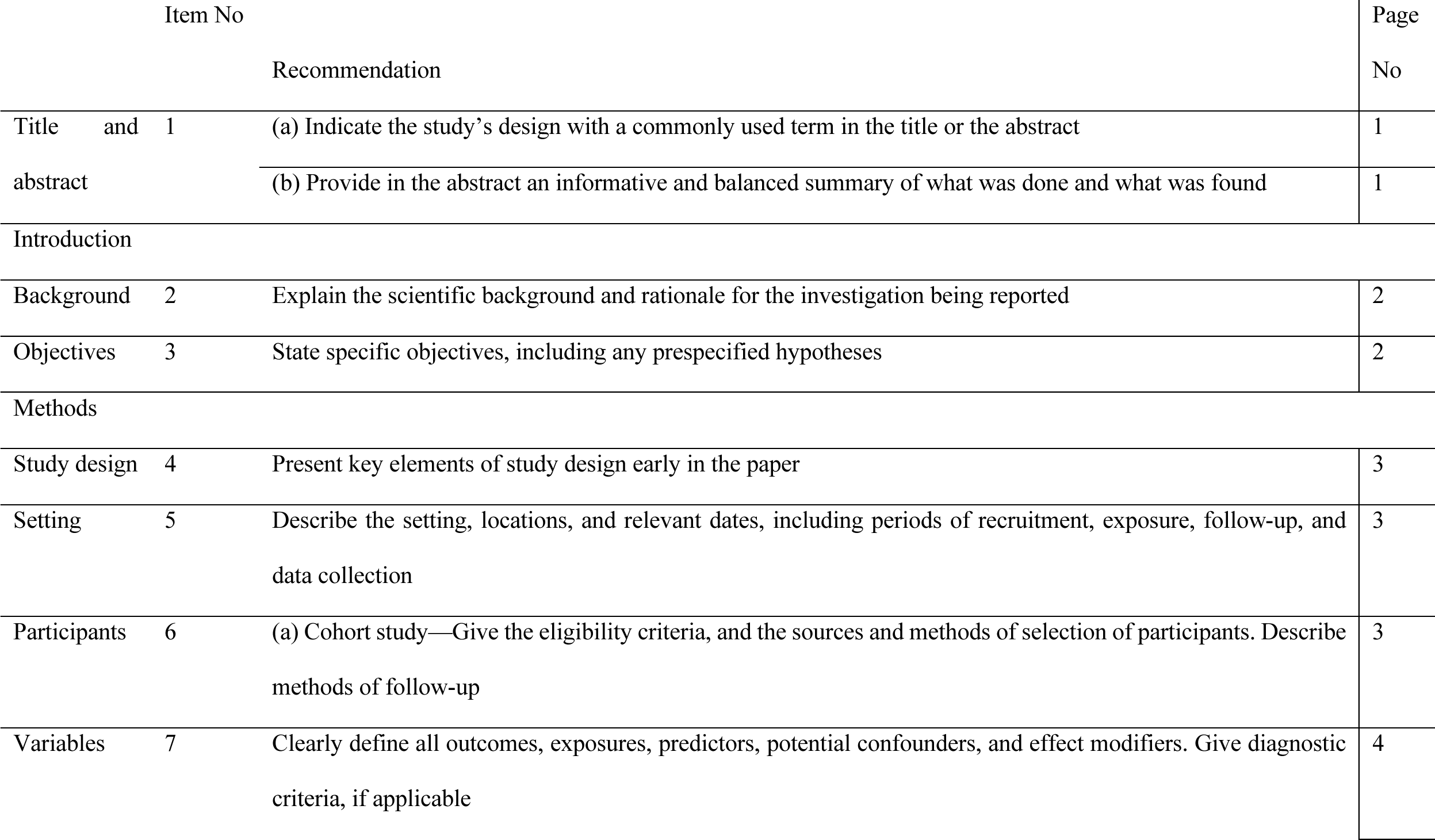
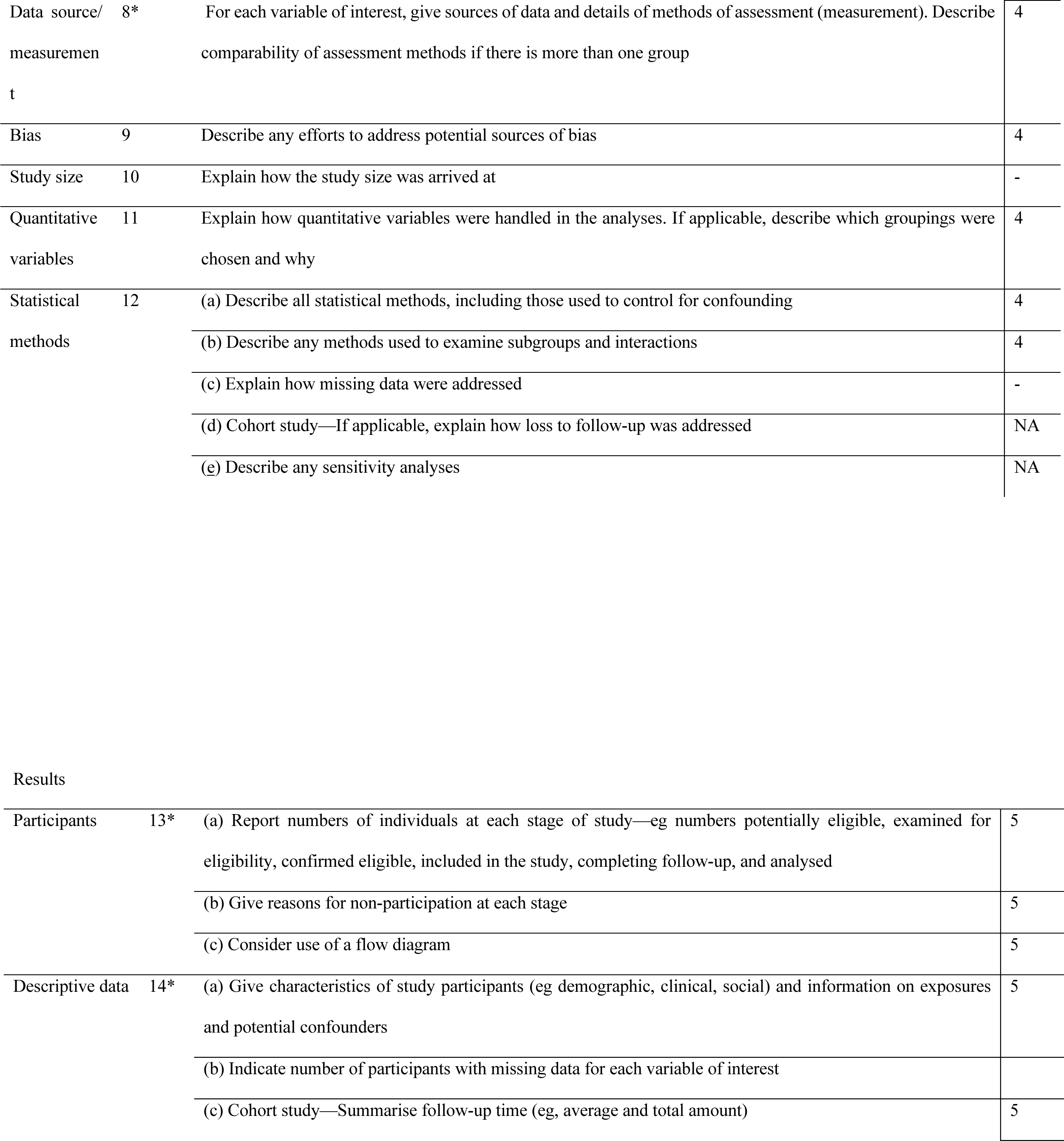
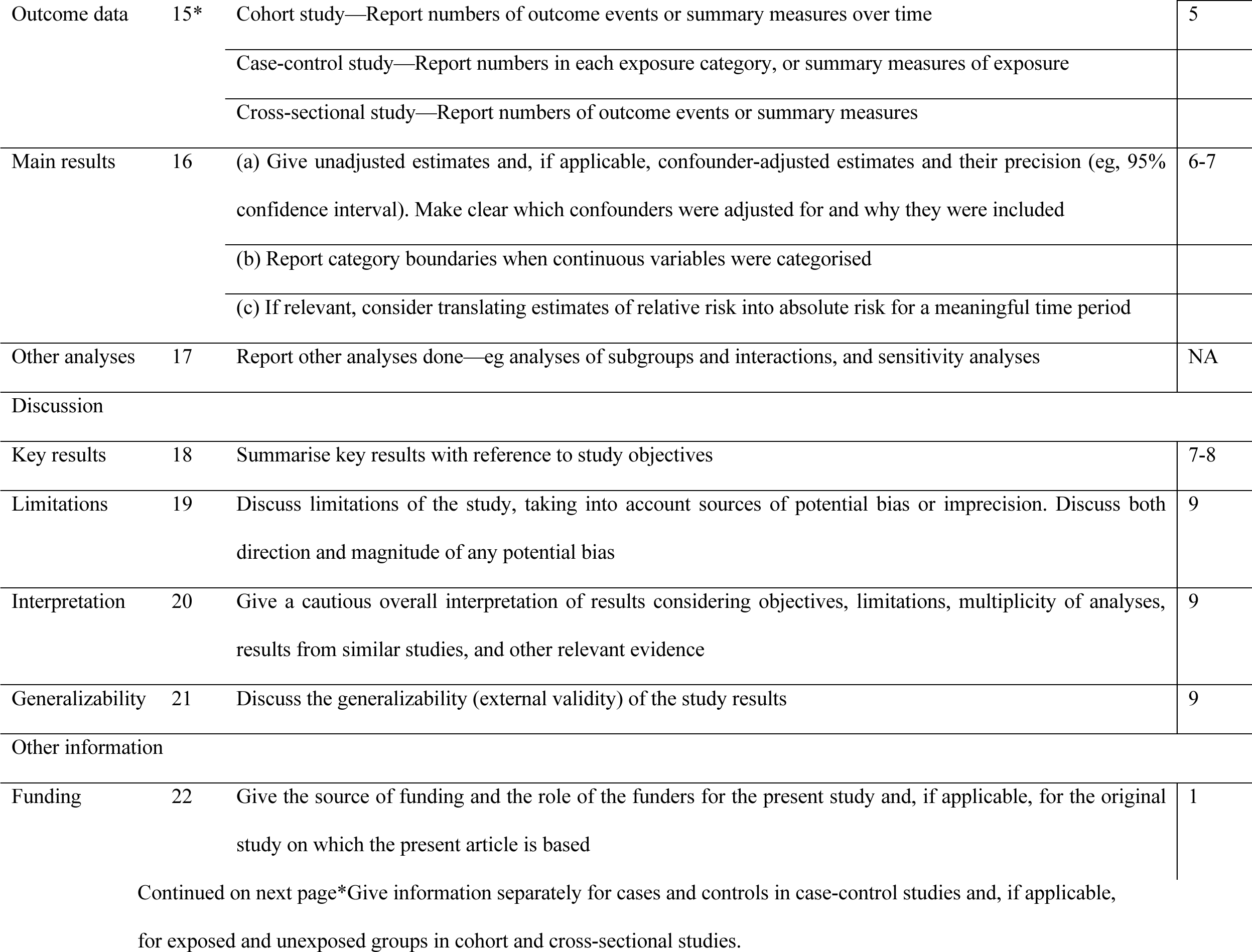
STROBE Statement—checklist of items for reporting of observational studies.

SUPPLEMENT 2: Social Vulnerability Index estimation 1. Social Vulnerability Index The SVI is derived from 16 social attributes, which measures overall vulnerability of a census tract and vulnerability across four thematic subdomains (1) Socioeconomic Status (below poverty, unemployed, income, no high school diploma), (2) Household composition and disability (aged ζ65 years, aged :< 17, disability, single-parent households), (3) Minority status and language (minority, speak English “less than well”), and (4) Housing type and transportation (multi-unit structures, mobile homes, crowding, no vehicle, group quarters such as worker dormitories, skilled nursing facilities, or college dorms).

## Notes

### Competing Interest Statement

The authors have declared no competing interest.

### Funding Statement

This study did not receive any funding

### Author Declarations

Ethics Committee /IRB: Committee on Clinical Investigations of Beth Israel Deaconess Medical Center waived ethical approval for this work.

## REFERENCES

1. Brindise E, Elkhatib I, Kuruvilla A, Silva R. Temporal Trends in Incidence and Outcomes of Acute Pancreatitis in Hospitalized Patients in the United States From 2002 to 2013. Pancreas. Feb 2019;48(2):169–175. doi:10.1097/MPA.0000000000001228

2. Gapp J, Hall AG, Walters RW, Jahann D, Kassim T, Reddymasu S. Trends and Outcomes of Hospitalizations Related to Acute Pancreatitis: Epidemiology From 2001 to 2014 in the United States. Pancreas. Apr 2019;48(4):548–554. doi:10.1097/MPA.0000000000001275

3. Peery AF, Crockett SD, Murphy CC, et al. Burden and Cost of Gastrointestinal, Liver, and Pancreatic Diseases in the United States: Update 2021. Gastroenterology. Feb 2022;162(2):621–644. doi:10.1053/j.gastro.2021.10.017

4. Li CL, Jiang M, Pan CQ, Li J, Xu LG. The global, regional, and national burden of acute pancreatitis in 204 countries and territories, 1990-2019. BMC Gastroenterol. Aug 25 2021;21(1):332. doi:10.1186/s12876-021-01906-2

5. Iannuzzi JP, King JA, Leong JH, et al. Global Incidence of Acute Pancreatitis Is Increasing Over Time: A Systematic Review and Meta-Analysis. Gastroenterology. Jan 2022;162(1):122–134. doi:10.1053/j.gastro.2021.09.043

6. Vege SS, DiMagno MJ, Forsmark CE, Martel M, Barkun AN. Initial Medical Treatment of Acute Pancreatitis: American Gastroenterological Association Institute Technical Review. Gastroenterology. Mar 2018;154(4):1103–1139. doi:10.1053/j.gastro.2018.01.031

7. Fabreau GE, Leung AA, Southern DA, et al. Sex, socioeconomic status, access to cardiac catheterization, and outcomes for acute coronary syndromes in the context of universal healthcare coverage. Circ Cardiovasc Qual Outcomes. Jul 2014;7(4):540–9. doi:10.1161/CIRCOUTCOMES.114.001021

8. Bacon SL, Bouchard A, Loucks EB, Lavoie KL. Individual-level socioeconomic status is associated with worse asthma morbidity in patients with asthma. Respir Res. Dec 17 2009;10(1):125. doi:10.1186/1465-9921-10-125

9. World Health O. A conceptual framework for action on the social determinants of health. Geneva: World Health Organization; 2010.

10. Braveman P, Gottlieb L. The social determinants of health: it’s time to consider the causes of the causes. Public Health Rep. Jan-Feb 2014;129 Suppl 2(Suppl 2):19-31. doi:10.1177/00333549141291S206

11. Centers for Disease Control and Prevention/ Agency for Toxic Substances and Disease Registry/ Geospatial Research, Analysis, and Services Program. CDC/ATSDR Social Vulnerability Index[2020] Database US https://www.atsdr.cdc.gov/placeandhealth/svi/index.html. (Last Accessed on January 31, 2024). .

12. von Elm E, Altman DG, Egger M, Pocock SJ, Gøtzsche PC, Vandenbroucke JP. Strengthening the Reporting of Observational Studies in Epidemiology (STROBE) statement: guidelines for reporting observational studies. Bmj. Oct 20 2007;335(7624):806-8. doi:10.1136/bmj.39335.541782.AD

13. International Classification of Diseases, Tenth Revision, Clinical Modification (ICD-10-CM); [updated 2017 Aug 18; reviewed 2017 Aug 18; cited 2017 Oct 11]. Available from: http://www.cdc.gov/nchs/icd/icd10cm.html (Last Accessed on January 31, 2024).

14. International Classification of Diseases NR, Clinical Modification (ICD-9-CM); [updated 2010 Sep 21; reviewed 2010 Sep 21; cited 2010 Nov 23]. Available from: http://www.cdc.gov/nchs/icd/icd9cm.htm. (Accessed on January 31, 2024).

15. Banks PA, Bollen TL, Dervenis C, et al. Classification of acute pancreatitis--2012: revision of the Atlanta classification and definitions by international consensus. Gut. Jan 2013;62(1):102–11. doi:10.1136/gutjnl-2012-302779

16. McHenry N, Shah I, Ahmed A, Freedman SD, Kothari DJ, Sheth SG. Racial Variations in Pain Management and Outcomes in Hospitalized Patients With Acute Pancreatitis. Pancreas. Oct 1 2022;51(9):1248–1250. doi:10.1097/MPA.0000000000002160

17. Anderson KL, Shah I, Tintara S, et al. Evaluating the Clinical Characteristics and Outcomes of Idiopathic Acute Pancreatitis: Comparison With Nonidiopathic Acute Pancreatitis Over a 10-Year Period. Pancreas. Oct 1 2022;51(9):1167–1170. doi:10.1097/MPA.0000000000002159

18. Charlson ME, Pompei P, Ales KL, MacKenzie CR. A new method of classifying prognostic comorbidity in longitudinal studies: development and validation. J Chronic Dis. 1987;40(5):373–83. doi:10.1016/0021-9681(87)90171-8

19. Wu BU, Johannes RS, Sun X, Tabak Y, Conwell DL, Banks PA. The early prediction of mortality in acute pancreatitis: a large population-based study. Gut. Dec 2008;57(12):1698–703. doi:10.1136/gut.2008.152702

20. Gao W, Yang HX, Ma CE. The Value of BISAP Score for Predicting Mortality and Severity in Acute Pancreatitis: A Systematic Review and Meta-Analysis. PLoS One. 2015;10(6):e0130412. doi:10.1371/journal.pone.0130412

21. Shaw GK, Waller S, Latham CJ, Dunn G, Thomson AD. The detoxification experience of alcoholic in-patients and predictors of outcome. Alcohol Alcohol. May-Jun 1998;33(3):291–303. doi:10.1093/oxfordjournals.alcalc.a008393

22. Livne O, Feinn R, Knox J, et al. Alcohol withdrawal in past-year drinkers with unhealthy alcohol use: Prevalence, characteristics, and correlates in a national epidemiologic survey. Alcohol Clin Exp Res. Mar 2022;46(3):422–433. doi:10.1111/acer.14781

23. Mulia N, Tam TW, Schmidt LA. Disparities in the use and quality of alcohol treatment services and some proposed solutions to narrow the gap. Psychiatr Serv. May 1 2014;65(5):626–33. doi:10.1176/appi.ps.201300188

24. Reoux JP, Miller K. Routine hospital alcohol detoxification practice compared to symptom triggered management with an Objective Withdrawal Scale (CIWA-Ar). Am J Addict. Spring 2000;9(2):135–44. doi:10.1080/10550490050173208

25. Social Deprivation Index (SDI) . Available at: https://www.graham-center.org/maps-data-tools/social-deprivation-index.html (Last Accessed on January 31.

26. Loney T, Nagelkerke NJ. The individualistic fallacy, ecological studies and instrumental variables: a causal interpretation. Emerg Themes Epidemiol. 2014;11:18. doi:10.1186/1742-7622-11-18

27. Greenland S, Morgenstern H. Ecological bias, confounding, and effect modification. Int J Epidemiol. Mar 1989;18(1):269–74. doi:10.1093/ije/18.1.269

28. Fazel S, Geddes JR, Kushel M. The health of homeless people in high-income countries: descriptive epidemiology, health consequences, and clinical and policy recommendations. Lancet. Oct 25 2014;384(9953):1529-40. doi:10.1016/S0140-6736(14)61132-6

29. Doran KM, Boyer AP, Raven MC. Health Care for People Experiencing Homelessness-What Outcomes Matter? JAMA Netw Open. Mar 1 2021;4(3):e213837. doi:10.1001/jamanetworkopen.2021.3837

30. Spielman SE, Tuccillo J, Folch DC, et al. Evaluating social vulnerability indicators: criteria and their application to the Social Vulnerability Index. Natural Hazards. 2020/01/01 2020;100(1):417-436. doi:10.1007/s11069-019-03820-z

31. Kandasamy C, Shah I, Yakah W, et al. The Impact of an Inpatient Pancreatitis Service and Educational Intervention Program on the Outcome of Acute Pancreatitis. Am J Med. Mar 2022;135(3):350–359 e2. doi:10.1016/j.amjmed.2021.09.021

